# Short-term outcomes associated with temozolomide vs. PCV chemotherapy for 1p/19q-codeleted WHO grade III anaplastic oligodendrogliomas: a national evaluation

**DOI:** 10.1101/2020.03.29.20046805

**Authors:** Nayan Lamba, Malia McAvoy, Vasileios K. Kavouridis, Timothy R. Smith, Mehdi Touat, David A. Reardon, J. Bryan Iorgulescu

**Author notes:** **Corresponding author:** J. Bryan Iorgulescu, M.D., Department of Pathology, Brigham and Women’s Hospital, Department of Medical Oncology, Dana-Farber Cancer Institute, Harvard Medical School, 75 Francis St., Boston, MA 02115. This work was presented in part at the 2019 annual meeting of the Society for Neuro-Oncology.

## Abstract

**PURPOSE:** The optimal chemotherapy regimen between temozolomide (TMZ) and procarbazine, lomustine, and vincristine (PCV) remains uncertain for newly-diagnosed anaplastic oligodendroglioma (AO). We therefore addressed this question using a national database.

**METHODS:** Patients newly-diagnosed with 1p/19q-codeleted W.H.O. grade III AO between 2010-2016 were identified from the National Cancer Database. Predictors of receiving first-line single-agent TMZ vs. multi-agent PCV were assessed by multivariable logistic regression. Overall survival (OS) was estimated by Kaplan-Meier techniques and evaluated by multivariable Cox regression.

**RESULTS:** 1,360 AO patients were identified: 74.5% (n=1,013) treated with TMZ, 9.6% (n=131) with PCV, and 15.9% (n=216) with no chemotherapy in the first-line setting. In multivariable logistic analysis, PCV utilization increased from 2010 to 2016 (OR=1.38/year, 95%CI: 1.22-1.56, p<0.001) and was less commonly utilized in privately insured patients (OR=0.38 vs. uninsured, 95%CI: 0.15-0.97, p=0.04). In survival analyses (33.1% reached endpoint), there was no difference in unadjusted OS between TMZ (5yr-OS 60.1%, 95%CI: 55.9-64.1) and PCV (5yr-OS 61.1%, 95%CI: 45.6-73.5; p=0.42). There remained no OS difference between TMZ and PCV in the 75.9% (n=1,032) of AO patients that also received radiotherapy (p=0.51), in the Cox regression analysis adjusted by age, extent of resection, and radiotherapy (TMZ vs. PCV HR=1.31, 95%CI: 0.83-2.08, p=0.24), and in subgroup analyses that incorporate KPS or *MGMT* status.

**CONCLUSIONS:** In a national database of AOs managed in the ‘real-world’ setting, there is no difference in the short-term mortality between first-line TMZ and PCV chemotherapy. These findings provide preliminary data while we await the long-term results from the CODEL trial.

## Introduction

Diffuse gliomas are a common primary intracranial tumor associated with significant morbidity and mortality.^1^ W.H.O. grade III anaplastic oligodendrogliomas (AOs) are a rare subset of diffuse gliomas, comprising about 6% of all gliomas, and, per the 2016 W.H.O. classification, are defined by the presence of both an *IDH* mutation and 1p/19q chromosomal codeletion.^2^ The 1p/19q chromosomal codeletion is a strong prognostic marker associated with increased responsiveness to chemotherapy and superior outcomes in patients with diffuse gliomas.^3,4^ However, despite recent advances in understanding its distinct molecular profile, the optimal therapeutic regimen for newly-diagnosed AOs remains unclear and controversial.^5^

The NCCN guidelines (version 3.2019) currently recommend maximal safe surgical resection followed by radiotherapy (RT) and chemotherapy for AOs.^4,6^ The addition of chemotherapy to RT in patients with AO stems from two major randomized controlled trials that demonstrated improved survival among patients harboring a 1p/19q-codeletion who received procarbazine, lomustine, and vincristine (PCV) chemotherapy with RT, as compared to those who received RT alone.^7,8^ More recently, in light of the results of the EORTC 26981–22981/NCIC CE3 trial, as well as evidence demonstrating that temozolomide (TMZ) may have a lower toxicity profile and more convenient administration than PCV, there has been interest in assessing whether TMZ might replace PCV in the treatment of AOs.^4^ The CODEL trial (NCT00887146), which presently randomizes newly-diagnosed oligodendroglioma patients to RT with either adjuvant PCV or with concomitant TMZ (followed by adjuvant TMZ), is expected to answer this question in the future;^9^ however, until its results become available, there is no clear consensus among the neuro-oncology community as to which regimen to use in the management of patients with AOs.^5^

Given the concerns surrounding PCV-related toxicity, we sought to compare outcomes for newly-diagnosed AO patients following TMZ or PCV in the first-line setting using a large national cancer database. As the results of the CODEL trial are years away, our study has the potential to offer clinical guidance about the efficacy of these regimens in the management of patients with newly-diagnosed AO.

## Materials and Methods

### Data Source and Study Design

The National Cancer Database (NCDB) is a nationwide database comprising more than 70% of newly diagnosed cancers in the United States.^10^ Patients were identified with histologically-confirmed newly-diagnosed diffuse gliomas between 2010 (1p/19q codeletion data are only incorporated in the NCDB as of 2010) and 2016, as defined by the ICD-O-3 histological codes (*i*.*e*. 9380-9382, 9400-9401, 9420, 9440-9442, 9450-9451, and 9460), a malignant behavior code (*i*.*e*. 3), and any brain site code (*i*.*e*. 71.0-71.9).^11^ Anaplastic oligodendrogliomas were defined herein using the primary brain-specific variables for W.H.O. grade III and for loss of heterozygosity/deletion of both chromosome arms 1p and 19q.^12^ A subset of cases with 1p/19q-codeletion were encoded in the NCDB as W.H.O. grade IV – likely reflecting the outmoded “glioblastoma with oligodendroglial features” – and herein were reclassified as W.H.O. grade III AOs in accordance with the revised 2016 W.H.O. classification. Patients were excluded if they had a prior diagnosis of cancer (thereby excluding W.H.O. grade II oligodendrogliomas that subsequently transformed into AO), were <20-years-old at diagnosis, were diagnosed at an index institution and treated entirely elsewhere, or if they were not 1p/19q-codeleted. Patients were also excluded if they lacked complete data regarding receipt of chemotherapy or RT.

### Variables and Statistical Analyses

Chemotherapy is encoded in the NCDB as single-agent or multi-agent, without further details. In order to confirm that single-agent referred to temozolomide and multi-agent to PCV, we reviewed all encoded chemotherapy data for AOs from registry-submitted data from two cancer centers (Brigham and Women’s Hospital/Dana-Farber Cancer Center and Massachusetts General Hospital) and validated their encoding using the medical records. Associations between clinicopathologic factors, including age at diagnosis, sex, race/ethnicity, Charlson-Deyo comorbidity index, year of diagnosis, insurance status, hospital type and location, tumor size (greatest dimension), extent of resection (EOR), and receipt of single-agent vs. multi-agent chemotherapy were assessed with multivariable logistic regression. Brain-specific EOR was stratified as biopsy-only, subtotal resection, or gross total resection. *IDH* mutation status is not encoded in the NCDB.

As previously described, overall survival (OS) was measured from the date of AO diagnosis to the date of death, with patients censored at the date of most recent follow-up.^12,13^ OS was estimated by Kaplan-Meier methods and compared by log-rank tests and Cox regression analysis. Multivariable Cox proportional hazards regression was used to evaluate the association between chemotherapy regimen and OS. Due to limited follow-up, the NCDB excludes OS data for patients diagnosed in the final year of the dataset, which for this dataset was 2016 – therefore AO patients diagnosed in 2016 are excluded from survival analyses. Because Karnofsky performance status (KPS, stratified as ≤60 vs. >60) and *MGMT* promoter methylation status were only encoded in a minority of patients (17.2% and 32.5% herein, respectively), these variables were separately added to the multivariable Cox regression analysis as subgroup analyses. STATA (v. 14.2, StataCorp) was used for statistical analyses, with 2-sided p-values <0.05 specified as significant. This study was approved by the Partners HealthCare institutional review board (#2015P002352).

## Results

1,360 U.S. patients with newly-diagnosed 1p/19q-codeleted W.H.O. grade III AOs were identified; of whom 74.5% (n=1,013) received single-agent chemotherapy, 9.6% (n=131) received multi-agent chemotherapy, and 15.9% (n=216) had no chemotherapy in the first-line setting. From two of the NCDB’s institutions, 73 1p/19q-codeleted AOs treated with chemotherapy between 2000-2017 were encoded and submitted to registries. Of these, 84.9% (n=62) and 15.1% (n=11) were encoded as first-line single-agent and multi-agent chemotherapy and were validated by medical chart review to represent TMZ and PCV, respectively, in all cases.

In multivariable logistic analysis of the national sample, privately insured patients were significantly less likely to receive multi-agent PCV (odds ratio [OR] 0.38, 95% confidence interval [CI]: 0.15-0.97, p=0.04) than uninsured patients (Table 1). AOs that were more recently diagnosed, however, were significantly associated with increased utilization of PCV (OR 1.38/year, 95%CI: 1.22-1.56, p<0.001). Receipt of single-agent TMZ vs. multi-agent PCV chemotherapy was otherwise independent of AO patients’ sex, age, race/ethnicity, comorbidity index, EOR, RT, tumor size (greatest dimension), hospital type, or hospital geographic location (all p>0.05, Table 1). 75.9% (n=1,032) of AO patients also received RT (42.7% as 60 Gy and 33.2% as 59.4 Gy). RT began a median of 36 days (interquartile range [IQR] 28-47) after definitive resection; and single-agent TMZ chemotherapy often began on the same day as RT (median difference 0 days, IQR: 0-0), whereas multi-agent PCV chemotherapy often began following RT (median difference 64 days, IQR: 0-78.5; t-test p<0.001).

**Table 1.**
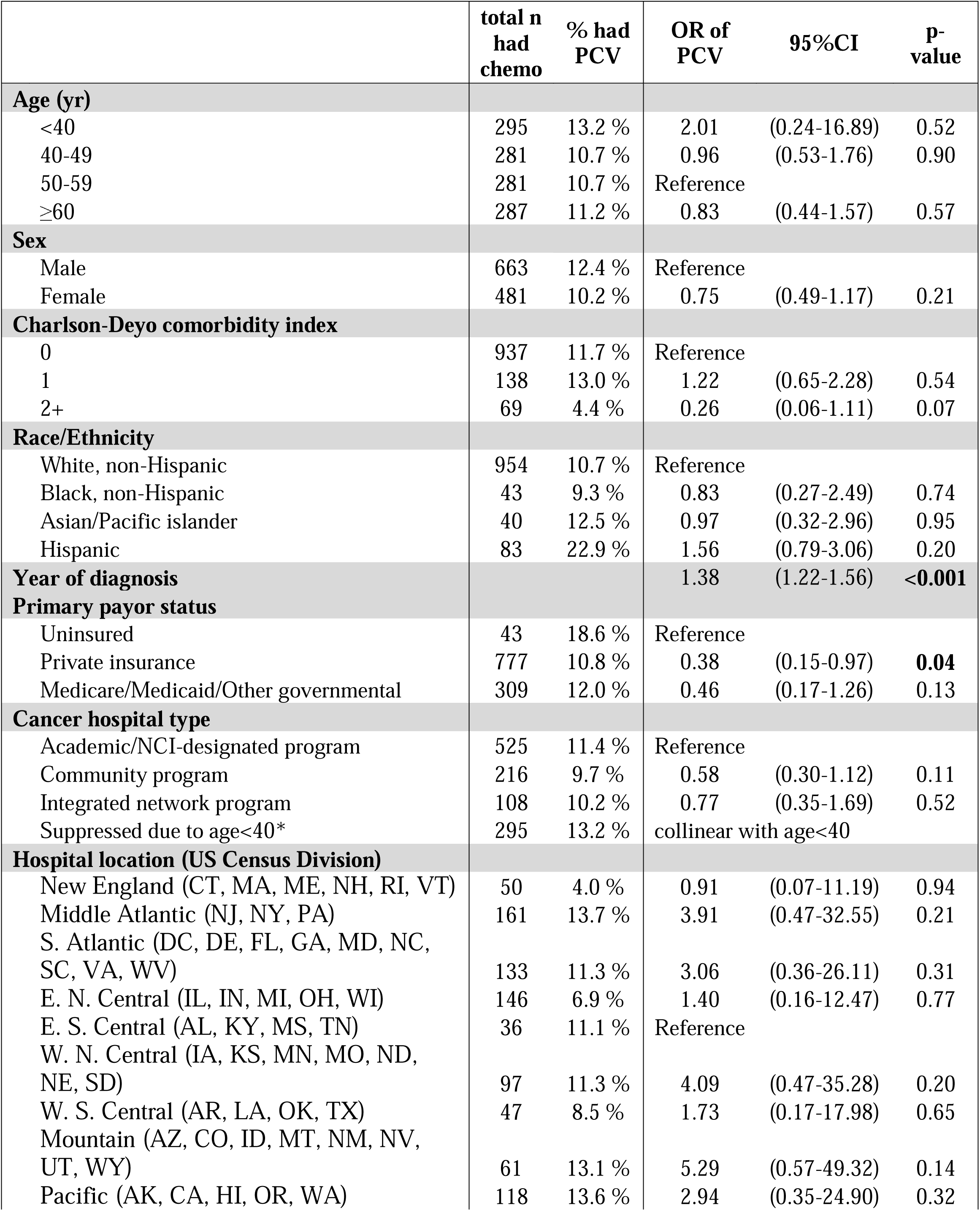
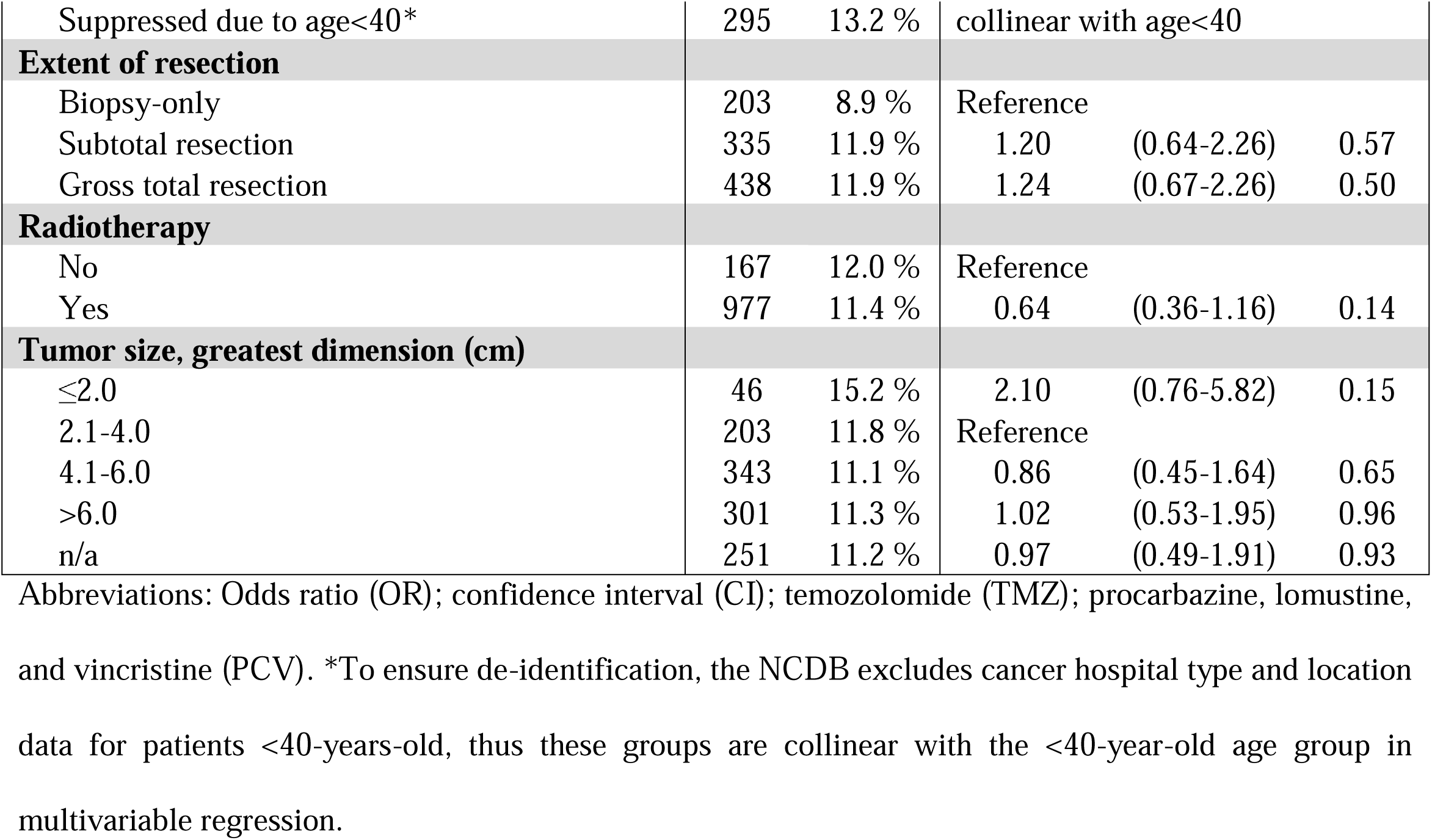
Multivariable logistic regression analysis of factors associated with utilization of PCV, as compared to TMZ.

In terms of OS, the median follow-up was 27.7 months (interquartile range [IQR] 12.4-49.9 months) with 33.1% of patients reaching the endpoint of mortality in the national sample. Unadjusted OS estimates were not significantly different between single-agent TMZ (5yr-OS 60.1%, 95%CI: 55.9-64.1) and multi-agent PCV chemotherapy (5yr-OS 61.1%, 95%CI: 45.6-73.5; Cox regression p=0.42), but both were significantly better than no chemotherapy (5yr-OS 46.5%, 95%CI: 36.8-55.7, p≤0.001; Fig. 1). In the 1,032 patients who also had RT, there remained no significant OS difference between TMZ (5yr-OS 56.1%, 95%CI: 51.4-60.6) and PCV (5yr-OS 54.0%, 95%CI: 36.0-68.9, p=0.51). In the 15.9% of AO patients who did not receive chemotherapy, 53.5% had resection alone, 19.8% had resection with RT, and 4.8% had biopsy with RT.

**Fig 1.**
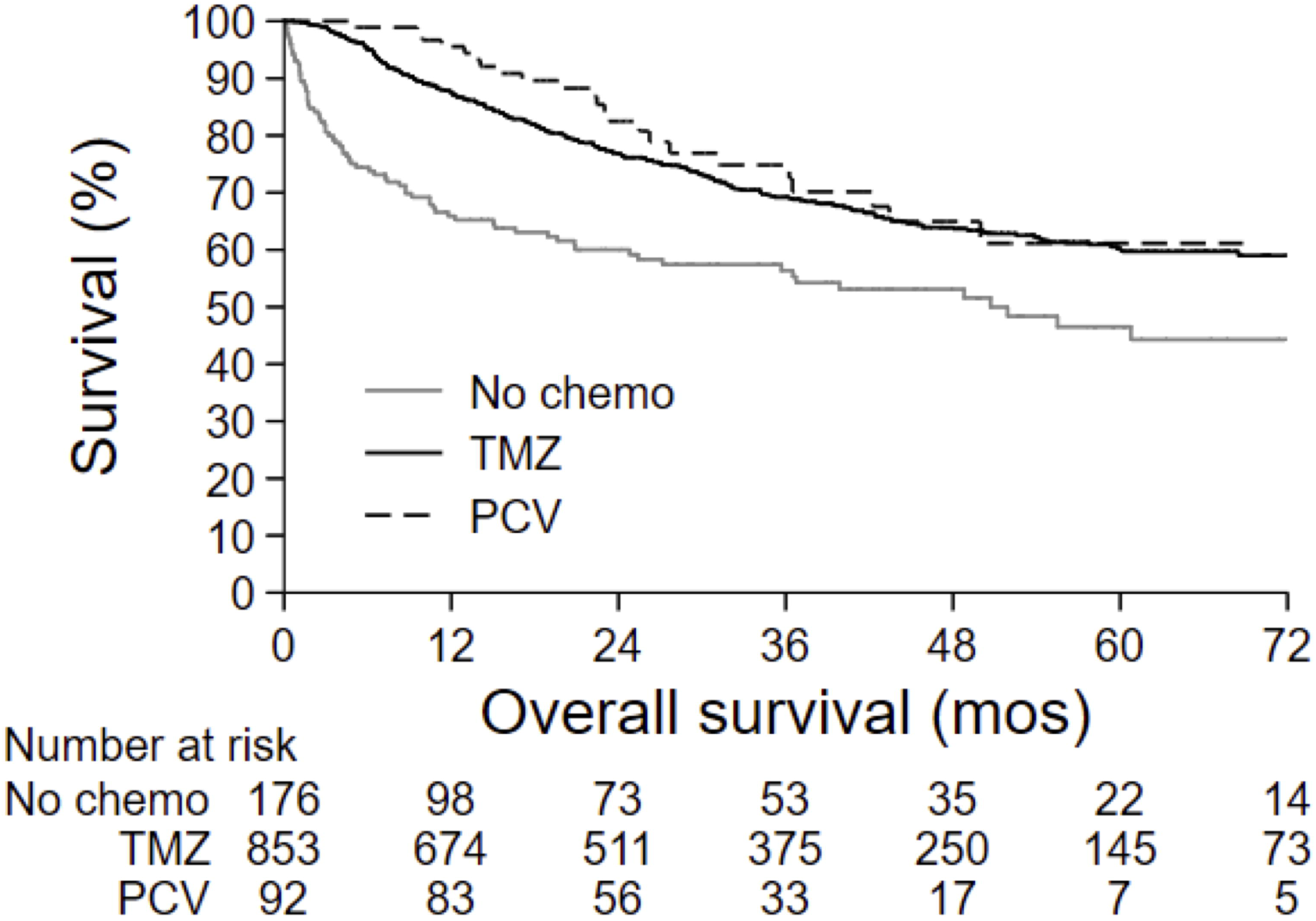
There is no difference in the unadjusted OS curves between single-agent TMZ (solid line) and multi-agent PCV (dashed line) treatment for AOs (Cox regression p=0.42)

In multivariable Cox regression analysis adjusted for age, EOR, and RT – factors previously shown to have prognostic value in AOs – there was no difference in OS between first-line TMZ (HR 1.31, 95%CI: 0.83-2.08, p=0.24) and PCV chemotherapy, and either regimen significantly improved OS compared to no chemotherapy (Table 2). The lack of an OS difference persisted in subgroup analyses that incorporated KPS (n=188, TMZ HR 0.92 vs. PCV, 95%CI: 0.12-6.94, p=0.94) and *MGMT* promoter methylation status (n=265, TMZ HR 1.04 vs. PCV, 95%CI: 0.50-2.16, p=0.91).

**Table 2.** Multivariable Cox regression analysis of OS in AOs.

|  | HR | 95%CI | p-value |
| --- | --- | --- | --- |
| **Chemotherapy** |  |  |  |
| None | Reference |  |  |
| TMZ | 0.51 | (0.38-0.68) | <0.001 |
| PCV | 0.39 | (0.23-0.65) | <0.001 |
| **Age (yr)** |  |  |  |
| <40 | 0.84 | (0.54-1.29) | 0.42 |
| 40-49 | Reference |  |  |
| 50-59 | 2.52 | (1.76-3.62) | <0.001 |
| 60+ | 4.19 | (2.96-5.93) | <0.001 |
| **Extent of resection** |  |  |  |
| Biopsy-only | Reference |  |  |
| Subtotal resection | 0.58 | (0.44-0.77) | <0.001 |
| Gross total resection | 0.60 | (0.46-0.78) | <0.001 |
| **Radiotherapy** |  |  |  |
| Yes (ref no) | 1.27 | (0.95-1.69) | 0.11 |

Abbreviations: Hazard ratio (HR); confidence interval (CI); temozolomide (TMZ); procarbazine, lomustine, and vincristine (PCV).

## Discussion

In this national analysis of 1,360 patients with newly-diagnosed 1p/19q-codeleted W.H.O. grade III anaplastic oligodendroglioma, we found no significant difference in short-term OS between patients treated with first-line single-agent (TMZ-based) vs. multi-agent (PCV-based) chemotherapy regimens. These findings persisted after adjusting for key prognostic variables of age at diagnosis, EOR, and RT, together providing support for the similarity of short-term OS outcomes for first-line TMZ vs. PCV in this patient population.

Two landmark trials established the chemosensitivity of 1p/19q-codeleted oligodendrogliomas and defined a role for PCV in the treatment of AOs: Radiation Therapy Oncology Group (RTOG) 9402 and the European Organization for Research and Treatment of Cancer (EORTC) 26951 both showed that among patients with 1p/19q-codeleted tumors, treatment with RT and PCV resulted in significant improvements in OS as compared to treatment with adjuvant RT alone.^7,8^ However, despite the compelling evidence in favor of adjuvant PCV, its use in clinical practice has been limited by its significant toxicity.^14^ Specifically, lomustine can cause bone marrow suppression and procarbazine can result in intractable nausea and skin reactions that require its discontinuation. In addition, vincristine is not thought to achieve appreciable penetration of the blood-brain-barrier, potentially limiting the overall efficacy of PCV in the treatment of an intracranial tumor.^15^ In light of patients’ poor tolerance of and ability to complete treatment with PCV, TMZ has also been explored as a potential alternative in the treatment of AOs.

Numerous retrospective studies have demonstrated that TMZ may be a safe and efficacious alternative for AO treatment, achieving similar response rates as chemotherapy with PCV. In a clinical efficacy trial of 20 patients with AO, TMZ was given for up to 24 weeks following surgical resection: results demonstrated a high response to TMZ, with clinical improvement reported in 60% of patients and only two reports of grade 3/4 toxicity.^16^ Similarly, another phase II study assessed the use of upfront TMZ in the treatment of newly-diagnosed AO and found a median progression-free survival (PFS) of 62 months, with limited high-grade toxicities despite a dose-intense schedule.^17^ Robust tumor responsiveness and well-managed toxicity following TMZ for AO have been consistently observed in retrospective studies.^18,19^ It should be noted, however, that none of the above studies included RT, so it is possible that response rates might have been even higher had RT been added to TMZ therapy, although toxicity profiles may also have been more severe.

Despite these reports suggesting an opportunity for TMZ in first-line treatment of AO, whether TMZ can outperform PCV remains uncertain. There have been a limited number of retrospective studies that have attempted to compare these two chemotherapy regimens head-to-head; ever fewer of which have incorporated the 2016 revised W.H.O. histomolecular definitions of AOs. In a large multi-institutional retrospective study by Lassman *et al*., in which only 89 1p/19q-codeleted AOs were compared by chemotherapy agent, PCV-alone demonstrated improved time-to-progression (median 7.6 months compared to 3.3 months in TMZ-alone, p=0.02), but no difference in OS (p=0.16).^20^ In the AOs that also received RT, there was no difference in either time-to-progression or OS for PCV vs. TMZ.^20^ In the Neuro-Oncology Working Group (NOA) phase III trial NOA-04, in which patients with anaplastic gliomas (only 69 with 1p/19q-codeletion) were randomized to upfront RT or upfront chemotherapy (either PCV or TMZ), researchers ultimately found that chemotherapy alone was not superior to RT.^21^ In the subset (n=33) of CpG island hypermethylated (CIMP) 1p/19q-codeleted cases, PCV-alone demonstrated improved PFS, but not OS, compared to TMZ-alone. Notably, although NOA-04 demonstrated better tumor control with PCV, it also demonstrated worse toxicity with PCV. Among the anaplastic glioma patients treated with upfront chemotherapy, treatment was discontinued in 33% of patients treated with PCV (n=54), all due to toxicity, while none of the TMZ-treated patients (n=53) required TMZ discontinuation due to toxicity.^22^ Although the Lassman *et al*. study and NOA-04 had moderate follow-up durations of approximately 10 years, it is highly likely that their data had not yet reached maturity and were underpowered with respect to the question of PCV vs. TMZ for AOs. Similar to these studies, our data only capture the short-term outcomes of PCV vs. TMZ for AOs, and it is possible that with much longer follow-up one regimen may prove superior at later landmarks. To rigorously answer this question the phase III CODEL trial (NCT00887146) has been redesigned as a randomized noninferiority comparison of RT either followed by adjuvant PCV or with concomitant TMZ followed by adjuvant TMZ for grade II and III newly-diagnosed 1p/19q-codeleted oligodendrogliomas, powered for PFS, and secondarily assessing OS, toxicity, and quality of life outcomes.

Because the final results of CODEL are likely years away, we evaluated the short-term OS outcomes associated with PCV or TMZ as first-line treatment in a contemporary national cohort of 1,360 patients with newly-diagnosed 1p/19q-codeleted AOs. Nationally, utilization of TMZ vs. PCV was significantly affected by insurance status: nearly twice as many uninsured AO patients received PCV as privately insured patients – suggesting that insurance-related financial barriers may pose challenges to TMZ access. Although prior work has suggested that TMZ may be surpassing PCV as the regimen-of-choice for gliomas given the favorable toxicity profiles of TMZ,^23^ in our analysis utilization of PCV actually increased for AOs from 2010 to 2016. This increase may reflect the timely publication of results from the RTOG 9402 and EORTC 26951 randomized controlled trials, which firmly established the survival benefit of adding PCV to RT for 1p/19q-codeleted gliomas. TMZ vs. PCV rates were otherwise independent of patient age, sex, comorbidity status, tumor size, hospital type and location, RT, and EOR. In line with the studies’ survival findings above, we found no difference in short-term OS between AOs treated with first-line PCV versus TMZ. This included in the setting of RT, as well as after multivariable adjustment for age, EOR, RT, *MGMT* promoter methylation status, and/or KPS.

## Limitations

Although our analysis encompasses a national dataset, the NCDB’s limited encoding of 1p/19q-codeletion status only as of 2010 constrained our follow-up to a median of 28 months. As a result, our findings only apply to the short-term outcomes of 1p/19q-codeleted AO patients and do not capture any delayed differences between PCV and TMZ. Based on a two-tailed log-rank power analysis using the Schoenfeld method, this sample set has 90% power to detect a 0.56 HR between the OS of TMZ and PCV. Although only results from a randomized trial comparing PCV and TMZ will be able to ultimately answer this question, our findings suggest that either PCV or TMZ may offer comparable early OS outcomes in the first-line management of newly-diagnosed AOs and support the tailoring of the TMZ vs. PCV selection to each patient’s unique circumstances.

Even though national registry-based databases like the NCDB provide an unparalleled look at outcomes in a real-life treatment population, they are constricted by several key limitations. First-line chemotherapy is encoded as single-agent and multi-agent, without details about agents, doses, early and late toxicities, or duration of treatment. To help address this limitation, registry-submitted data for AOs from two institutions were evaluated, in which all single-agent cases were TMZ and all multi-agent cases were PCV. Furthermore, only the initial courses of therapy are encoded in the NCDB, so it is possible that some cases received additional single-agent or multi-agent chemotherapy as second-line therapy. Molecular data are also very limited in registry data: 1p/19q-codeletion status was only incorporated as of 2010, resulting in a median follow-up for this analysis of 28 months, without details about the diagnostic methodology. *IDH* mutational status is presently lacking in registry data, so all oligodendrogliomas herein were defined by their 1p/19q-codeletion and thus were also likely *IDH* mutant.^11^ *MGMT* promoter methylation was encoded in a minority of cases, but CpG island hypermethylation data are not included. Additionally, only OS is encoded for outcomes, precluding the evaluation of PFS for PCV vs. TMZ in AOs.

## Conclusions

In this study of 1,360 patients with newly-diagnosed 1p/19q-codeleted AO, we found no difference in the short-term OS between first-line multi-agent PCV and single-agent TMZ chemotherapy, including in the setting of RT. In addition, we found overall utilization of PCV to be on the rise between 2010 and 2016, and that uninsured patients were significantly more likely to receive PCV than privately insured patients – suggesting that there may exist insurance-related financial barriers to the access of TMZ. While results from the CODEL trial will ultimately offer guidance with respect to optimal management for patients with newly-diagnosed AO, our results offer preliminary evidence that both TMZ and PCV chemotherapy regimens may be viable options in the first-line setting.

## Data Availability

Availability of data and material: Data are publicly available by application to the American College of Surgeons in accordance with the National Cancer Database’s data use agreement. Code availability: Available upon request.

## Acknowledgements

The authors are grateful to the cancer registrars at BWH/DFCI and MGH for their invaluable assistance in data acquisition. JBI acknowledges support by the NIH (T32HL007627).

## Declarations

The authors have no declarations.

## Availability of data and material

Data are publicly available by application to the American College of Surgeons in accordance with the National Cancer Database’s data use agreement.

## Code availability

Available upon request.

## Authors’ contributions

Conception and study design: JBI. Data collection: NL, MM, VK. Data analysis: JBI. Data interpretation and Manuscript writing: All authors. Critical review and revisions: All authors.

## References

1. Ostrom QT, Gittleman H, Liao P, et al. CBTRUS Statistical Report: Primary brain and other central nervous system tumors diagnosed in the United States in 2010-2014. Neuro-Oncol. 2017. doi:10.1093/neuonc/nox158

2. Louis DN, Ohgaki H, Cavenee WK. World Health Organization Histological Classification of Tumours of the Central Nervous System. Vol 1. revised 4th. France: International Agency for Research on Cancer; 2016.

3. Cairncross JG, Ueki K, Zlatescu MC, et al. Specific genetic predictors of chemotherapeutic response and survival in patients with anaplastic oligodendrogliomas. J Natl Cancer Inst. 1998. doi:10.1093/jnci/90.19.1473

4. Omuro A, DeAngelis LM. Glioblastoma and other malignant gliomas: A clinical review. JAMA. 2013. doi:10.1001/jama.2013.280319

5. Ruff MW, Buckner JC, Johnson DR, van den Bent MJ, Geurts M. Neuro-Oncology Clinical Debate: PCV or temozolomide in combination with radiation for newly diagnosed high-grade oligodendroglioma. Neuro-Oncol Pract. 2019;6(1):17–21. doi:10.1093/nop/npy044

6. NCCN Clinical Practice Guidelines in Oncology: Central Nervous System Cancers. version 3.2019 (November 11,2019) https://www.nccn.org/professionals/physician_gls/pdf/cns_blocks.pdf.

7. Van Den Bent MJ, Brandes AA, Taphoorn MJB, et al. Adjuvant procarbazine, lomustine, and vincristine chemotherapy in newly diagnosed anaplastic oligodendroglioma: Long-term follow-up of EORTC brain tumor group study 26951. J Clin Oncol. 2013. doi:10.1200/JCO.2012.43.2229

8. Cairncross G, Wang M, Shaw E, et al. Phase III trial of chemoradiotherapy for anaplastic oligodendroglioma: Long-term results of RTOG 9402. J Clin Oncol. 2013. doi:10.1200/JCO.2012.43.2674

9. Weller M, van den Bent M, Tonn JC, et al. European Association for Neuro-Oncology (EANO) guideline on the diagnosis and treatment of adult astrocytic and oligodendroglial gliomas. Lancet Oncol. 2017. doi:10.1016/S1470-2045(17)30194-8

10. Boffa DJ, Rosen JE, Mallin K, et al. Using the National Cancer Database for Outcomes Research: A Review. JAMA Oncol. 2017;3(12):1722–1728. doi:10.1001/jamaoncol.2016.6905

11. Iorgulescu JB, Torre M, Harary M, et al. The Misclassification of Diffuse Gliomas: Rates and Outcomes. Clin Cancer Res. 2019;25(8):2656–2663. doi:10.1158/1078-0432.CCR-18-3101

12. Harary M, Kavouridis VK, Torre M, et al. Predictors and early survival outcomes of maximal resection in WHO grade II 1p/19q-codeleted oligodendrogliomas. Neuro-Oncol. September 2019. doi:10.1093/neuonc/noz168

13. Iorgulescu JB, Harary M, Zogg CK, et al. Improved Risk-Adjusted Survival for Melanoma Brain Metastases in the Era of Checkpoint Blockade Immunotherapies: Results from a National Cohort. Cancer Immunol Res. July 2018. doi:10.1158/2326-6066.CIR-18-0067

14. Ruff MW, Buckner JC, Johnson DR, Van Den Bent MJ, Geurts M. Neuro-oncology clinical debate: Pcv or temozolomide in combination with radiation for newly diagnosed high-grade oligodendroglioma. Neuro-Oncol Pract. 2019. doi:10.1093/nop/npy044

15. Boyle FM, Eller SL, Grossman SA. Penetration of intra-arterially administered vincristine in experimental brain tumor. Neuro-Oncol. 2004. doi:10.1215/s1152851703000516

16. Taliansky-Aronov A, Bokstein F, Lavon I, Siegal T. Temozolomide treatment for newly diagnosed anaplastic oligodendrogliomas: A clinical efficacy trial. J Neurooncol. 2006. doi:10.1007/s11060-005-9020-1

17. Ahluwalia MS, Xie H, Dahiya S, et al. Efficacy and patient-reported outcomes with dose-intense temozolomide in patients with newly diagnosed pure and mixed anaplastic oligodendroglioma: a phase II multicenter study. J Neurooncol. 2015. doi:10.1007/s11060-014-1684-y

18. Mikkelsen T, Doyle T, Anderson J, et al. Temozolomide single-agent chemotherapy for newly diagnosed anaplastic oligodendroglioma. J Neurooncol. 2009. doi:10.1007/s11060-008-9735-x

19. Ducray F, Sierra Del Rio M, Carpentier C, et al. Up-front temozolomide in elderly patients with anaplastic oligodendroglioma and oligoastrocytoma. J Neurooncol. 2011. doi:10.1007/s11060-010-0264-z

20. Lassman AB, Iwamoto FM, Cloughesy TF, et al. International retrospective study of over 1000 adults with anaplastic oligodendroglial tumors. Neuro-Oncol. 2011. doi:10.1093/neuonc/nor040

21. Wick W, Society for the NWG (NOA) of the GC, Roth P, et al. Long-term analysis of the NOA-04 randomized phase III trial of sequential radiochemotherapy of anaplastic glioma with PCV or temozolomide. Neuro-Oncol. 2016.

22. Wick W, Hartmann C, Engel C, et al. NOA-04 randomized phase III trial of sequential radiochemotherapy of anaplastic glioma with Procarbazine, Lomustine, and Vincristine or Temozolomide. J Clin Oncol. 2009. doi:10.1200/JCO.2009.23.6497

23. Yu T, Kang HC, Lim DH, et al. Pattern of care of anaplastic oligodendroglioma and oligoastrocytoma in a Korean population: the Korean radiation oncology group study 13-12. J Neurooncol. 2015. doi:10.1007/s11060-014-1660-6

